# Feature-Based Parametric Response Mapping on Thoracic Computed Tomography for Robust Disease Classification in COPD

**DOI:** 10.64898/2026.04.24.26351675

**Authors:** Ali Namvar, Bingzhao Shan, Benjamin Hoff, Wassim W. Labaki, Susan Murray, Alexander J. Bell, Stefanie Galban, Ella A. Kazerooni, Fernando J. Martinez, Charles R. Hatt, MeiLan K. Han, Craig J. Galban, Sundaresh Ram

## Abstract

**Purpose:** To develop an interpretable feature-based Deep Parametric Response Mapping (PRM_D_) method that combines wavelet scattering convolution networks and machine learning to spatially detect and quantify functional small airways disease (fSAD) and emphysema on paired inspiratory-expiratory CT scans, with enhanced noise robustness.

**Materials and Methods:** In this retrospective analysis of prospectively acquired data (2007–2017), we developed and validated a deep learning-based PRM approach using paired CT scans from 8,972 tobacco-exposed COPDGene participants (≥10 pack-years; mean age 60.1 ± 8.8 years; 46.5% women), including controls with normal spirometry (n = 3,872; controls), PRISm (n = 1,089), GOLD 1–4 COPD (n = 4,011). Data were stratified into training, validation, and testing sets (24:6:70). PRM_D_ extracts translation-invariant image features using a wavelet scattering network and applies a subspace learning classifier to classify voxels as emphysema or non-emphysematous air trapping (fSAD). PRM_D_ was compared with conventional density-based PRM for voxel-wise agreement, correlation with pulmonary function, robustness to noise, and sensitivity to misregistration using Pearson correlation, Bland-Altman analysis, and paired t tests.

**Results:** PRM_D_ achieved 95% voxel-wise agreement with standard PRM (r = 0.98) while demonstrating significantly greater robustness under noise. PRM_D_ showed stronger correlations with FEV (emphysema: r = –0.54; fSAD: r = –0.51; *P* < 0.0001) than standard PRM (r = –0.42 for both; *P* < 0.0001). Under simulated high-noise conditions, standard PRM overestimated disease by ∼15%, whereas PRM_D_ limited error to < 5% (*P* < 0.001).

**Conclusion:** PRM_D_ provides an interpretable, feature-driven and noise-resilient alternative to traditional PRM for emphysema and fSAD classification, enhancing the reliability of CT-based COPD phenotyping for multi-center studies and low-dose imaging applications.

**Key Points:**

- This study introduces combined wavelet scattering and subspace learning for medical image segmentation, enabling accurate, interpretable voxel-level classification of emphysema and functional small airways disease on paired CT scans.
- The proposed Deep Parametric Response Mapping method demonstrated 95% voxel-wise agreement with standard Parametric Response Mapping and stronger correlations with spirometric measures, enhancing the clinical relevance of CT-based phenotyping for Chronic Obstructive Pulmonary Disease.
- Deep Parametric Response Mapping significantly improved robustness to image noise—reducing overestimation of emphysema and functional small airways disease from ∼15% to <5% (*P* < 0.001)—and benefits from reduced data requirements due to the fixed, mathematically defined filters used in wavelet scattering.

**Summary Statement:** Deep Parametric Response Mapping improves the accuracy and noise robustness of CT-based classification of emphysema and functional small airways disease using feature-based representations, enhancing the reliability of COPD phenotyping.

## Introduction

Chronic Obstructive Pulmonary Disease (COPD) is characterized by a combination of small airways disease and emphysema, leading to airflow limitation (1). Chest computed tomography (CT) imaging is widely used in COPD research and can quantitatively assess these disease components. In particular, parametric response mapping (PRM) is an established voxel-wise CT analysis technique that classifies lung tissue as normal, emphysema, or functional small airways disease (fSAD) by comparing paired inspiratory and expiratory scans (2, 3). PRM-derived measures of emphysema and fSAD have been shown to correlate with clinical indices and outcomes in COPD (2).

Nevertheless, the standard PRM technique relies on fixed density thresholds and image registration, which can make it sensitive to variations in CT image noise, image reconstruction settings, scanner types, and imperfect alignment of inspiratory/expiratory scans (4). Adaptive threshold methods, such as the Personalized Threshold Method (PTM), aim to mitigate these challenges by calculating a patient-specific cutoff for air trapping based on inspiratory lung density. While these methods improve air-trapping quantification, they still depend on intensity thresholding and may not fully overcome noise sensitivity (5). An alternative strategy is to leverage advanced image features and machine learning to recognize patterns and classify voxels, rather than using fixed thresholds (6).

In this study, we propose a feature-based deep parametric response mapping (PRM_D_) method that uses a wavelet scattering convolutional network (7, 8) to extract multi-scale texture features from lung CT images, combined with supervised subspace learning, to classify lung voxels into normal, fSAD, or emphysema. The wavelet scattering transform (a form of deep convolutional network with fixed filters) generates features that are translation invariant and robust to noise and deformations, potentially addressing the limitations of threshold-based PRM. We applied PRM_D_ to COPD subject CT scans from the COPDGene study and compared its outputs to conventional density-based PRM. We also evaluated the method’s performance across different disease severities and tested its stability under simulated noise. Our overall goal is to improve the reliability of CT-based COPD phenotyping by reducing sensitivity to technical factors, thereby paving the way for more consistent imaging biomarkers of COPD.

## Materials and Methods

### Study Population and Imaging

This analysis used anonymized thoracic CT scans and clinical data from the NIH-funded COPDGene^®^ cohort (9), a large multi-center longitudinal study of individuals with ≥10 pack-years tobacco exposure. Baseline characterization included symptoms, spirometry, and quantitative imaging as described previously (9). COPD was defined by post-bronchodilator FEV /FVC <0.70 per Global Initiative for Chronic Obstructive Lung Disease (GOLD) criteria (10). Demographics are summarized in Table 1.

**Table 1.** Baseline Summary of COPDGene Cohort, Highlighting Demographic, Spirometry, and Imaging Data for Training, Validation, Testing and Overall (N = 8972*)

| CHARACTERISTIC | TRAINING<br>(N = 2152) | VALIDATION<br>(N = 538) | TESTING<br>(N = 6282) | ALL<br>(N = 8972) |
| --- | --- | --- | --- | --- |
| Age (years) | 60.2 (8.5) | 59.9 (9.1) | 60.1 (8.9) | 60.1 (8.8) |
| Gender, N (% female) | 938 (43.6) | 248 (46.1) | 2982 (47.5) | 4168 (46.5) |
| Race, N (% Black) | 893 (41.5) | 193 (35.9) | 1798 (28.6) | 2884 (32.1) |
| BMI (kg/m<sup>2</sup>) | 29.1 (6.4) | 28.5 (6.8) | 28.2 (6.9) | 28.6 (6.7) |
| Currently Smoking, N (%) | 1028 (47.8) | 219 (40.7) | 3482 (55.4) | 4729 (52.7) |
| Smoking Pack Years | 45.1 (24.2) | 44.6 (24.7) | 45.7 (23.8) | 45.1 (24.1) |
| Age Smoking Initiated (years) | 16.4 (4.5) | 16.7 (4.1) | 16.5 (4.3) | 16.5 (4.3) |
| St. George's Respiratory Questionnaire Score (SGRQ) | 29.9 (21.1) | 30.3 (20.5) | 30.5 (20.2) | 30.2 (20.6) |
| GOLD Spirometric Stage (PRISm/GOLD 0/1/2/3/4) | 261/929/170/<br>415/249/128 | 65/232/43/<br>104/62/32 | 763/2711/498/<br>1208/728/374 | 1089/3872/711/<br>1727/1039/534 |
| FEV<sub>1</sub> (L) | 2.1 (0.6) | 2.2 (0.4) | 2.0 (0.7) | 2.1 (0.6) |
| FEV<sub>1</sub> % Predicted | 70.7 (12.6) | 69.7 (11.3) | 72.1 (12.6) | 70.8 (12.1) |
| FVC (L) | 3.2 (0.8) | 3.1 (0.6) | 3.3 (0.6) | 3.2 (0.7) |
| FVC % Predicted | 81.3 (12.9) | 80.9 (13.7) | 82.5 (13.4) | 81.6 (13.3) |
| FEV<sub>1</sub>/FVC | 0.7 (0.1) | 0.6 (0.1) | 0.7 (0.1) | 0.7 (0.1) |
| TLC (L) | 5.4 (1.3) | 5.3 (1.5) | 5.3 (1.4) | 5.3 (1.4) |
| FRC (L) | 3.3 (0.8) | 3.4 (1.0) | 3.4 (0.9) | 3.4 (0.9) |
| Mean HU Inspiration | -828.8 (43.3) | -830.2 (37.9) | -827.4 (41.5) | -828.8 (40.9) |
| **Mean HU Expiration** | -717.2 (54.6) | -723.5 (56.7) | -720.7 (52.1) | -720.5 (54.5) |
| **Segmental Wall Area %** | 52.6 (7.8) | 54.3 (8.2) | 53.5 (8.1) | 53.4 (8.0) |
| **Pi10** | 2.5 (0.6) | 2.6 (0.5) | 2.5 (0.7) | 2.5 (0.6) |
| **Emph-LAA %** | 6.2 (6.0) | 7.1 (6.7) | 6.8 (5.9) | 6.7 (6.2) |
| **AT-LAA %** | 23.1 (15.9) | 24.6 (14.8) | 23.8 (15.0) | 23.8 (15.2) |

NOTE: All values expressed as mean (SD) except categorical variables expressed as N (%). SD = standard deviation. GOLD = Global Initiative for Chronic Obstructive Lung Disease. FEV<sub>1</sub> = forced expiratory volume in the first second. FVC = forced vital capacity. TLC = total lung capacity measured on volumetric computed tomography of the chest at maximal inhalation. FRC = functional residual capacity measured on volumetric computed tomography of the chest at the end of tidal exhalation. LAA = low attenuation areas. Pi10 = square root of the wall area of a theoretical airway of 10 mm luminal perimeter, Emph = emphysema, AT = air trapping.
\*Due to missing data, age, BMI, smoking start age, sex, smoking status, exacerbation history, race and chronic bronchitis N=8,971; SGRQ N=8,970; pack years N=8,969; FEV<sub>1</sub>, FEV<sub>1</sub>% predicted, FVC, FVC% predicted, FEV<sub>1</sub>/FVC N=8,968; mMRC N=8,960; mean inspiratory HU and Pi10 N=8,959; wall area percent N=8,958; FEV<sub>1</sub> and FVC bronchodilator reversibility, N=8,860; FRC and mean expiratory HU N=8,042.

We analyzed baseline COPDGene data with paired inspiratory and expiratory CT scans at total lung capacity (TLC) and functional residual capacity (FRC), respectively, acquired via multi-detector CT scanners under standardized protocols (9). Subjects lacking complete paired CTs or with severe artifacts were excluded. The final cohort included 8,972 participants: normal spirometry (N=3,872), PRISm (N=1,089), GOLD 1-2 (N=2,438), and GOLD 3-4 (N=1,573). Mean age was 60.1 ± 8.8 years; 46.5% were female. Additional data included demographics, smoking history, respiratory symptoms, spirometric lung function, and quantitative CT metrics such as mean attenuation, lung volume, wall area percent, and pi10.

### Standard Density-based PRM Classification (Reference Standard)

We implemented conventional density threshold-based PRM as the reference standard for voxel classification. Following definitions from previous literature (2), lung voxels were classified as emphysema if inspiratory CT attenuation was < −950 Hounsfield Units (HU); functional small airways disease (fSAD) if inspiratory attenuation was ≥ −950 HU and expiratory attenuation < −856 HU; and normal if expiratory attenuation was ≥ −856 HU (11, 12). In practice, after image registration, we examined each voxel’s intensity in the aligned inspiratory and expiratory images to assign it to one of the three classes. For each subject, we also calculated the percentage of lung volume (number of voxels) falling into emphysema and fSAD categories as global summary metrics. These PRM-derived percentages served as the “ground truth” measurements of disease extent for subsequent comparisons.

### Deep PRM Method (Wavelet Scattering and Subspace Learning-Based Classification) Feature Extraction

The PRM_D_ method applies a 3D wavelet scattering transform to extract multi-scale, translation-invariant features from paired inspiratory and expiratory CT images. Wavelet scattering transforms are convolutional cascades using fixed (non-learned) filters followed by modulus and averaging operations (13). These features are robust to noise, stable under small deformations, and preserve high-frequency information while exhibiting fast energy decay (6, 8, 14–17).

We applied the transform independently to inspiratory and expiratory CT scans after image registration. Morlet wavelets (Gaussian-modulated sinusoids) were used as the mother wavelet. Our implementation used three spatial scales and 12 orientations, designed to capture texture structures ranging from fine (∼1–2 mm) to coarse (∼8–16 mm) resolution. After convolution, the modulus of each filter response was computed, followed by local averaging. This was repeated up to the second scattering order, producing scattering coefficients for each voxel in each scan. Scattering features from the same voxel in both scans were concatenated to form its final feature vector, encoding local lung texture across respiratory phases.

### Classification

Voxel-wise classification was performed using a supervised generative classifier based on affine subspace models trained via generalized principal component analysis (18). Two classifiers were trained independently: one for classifying emphysema using inspiratory CT features, and another for identifying air trapping using expiratory CT features. Ground-truth labels were derived from standard density-based PRM using fixed attenuation thresholds as described earlier.

The classifiers learned to model normal, emphysematous, and air-trapped lung tissue as separate affine spaces in the scattering feature domain. Class assignment was determined by projection error relative to each class-specific subspace. Classifiers were trained on a subset of the COPDGene dataset (described under Validation), minimizing misclassification error while avoiding overfitting.

### Post-Processing

Functional small airways disease (fSAD) was identified by comparing voxel-wise predictions from the two classifiers. Voxels classified as air trapping on expiratory CT but not emphysema on inspiratory CT were labeled as fSAD. Image registration ensured spatial correspondence between inspiratory and expiratory segmentations.

By leveraging fixed filters and a modular classification pipeline, PRM_D_ provides a deep, interpretable, and noise-robust approach to COPD phenotyping. The use of fixed scattering filters reduces training data requirements while enhancing model generalizability. A schematic representation of our proposed PRM_D_ method is shown in Figure 1.

**Figure 1.**
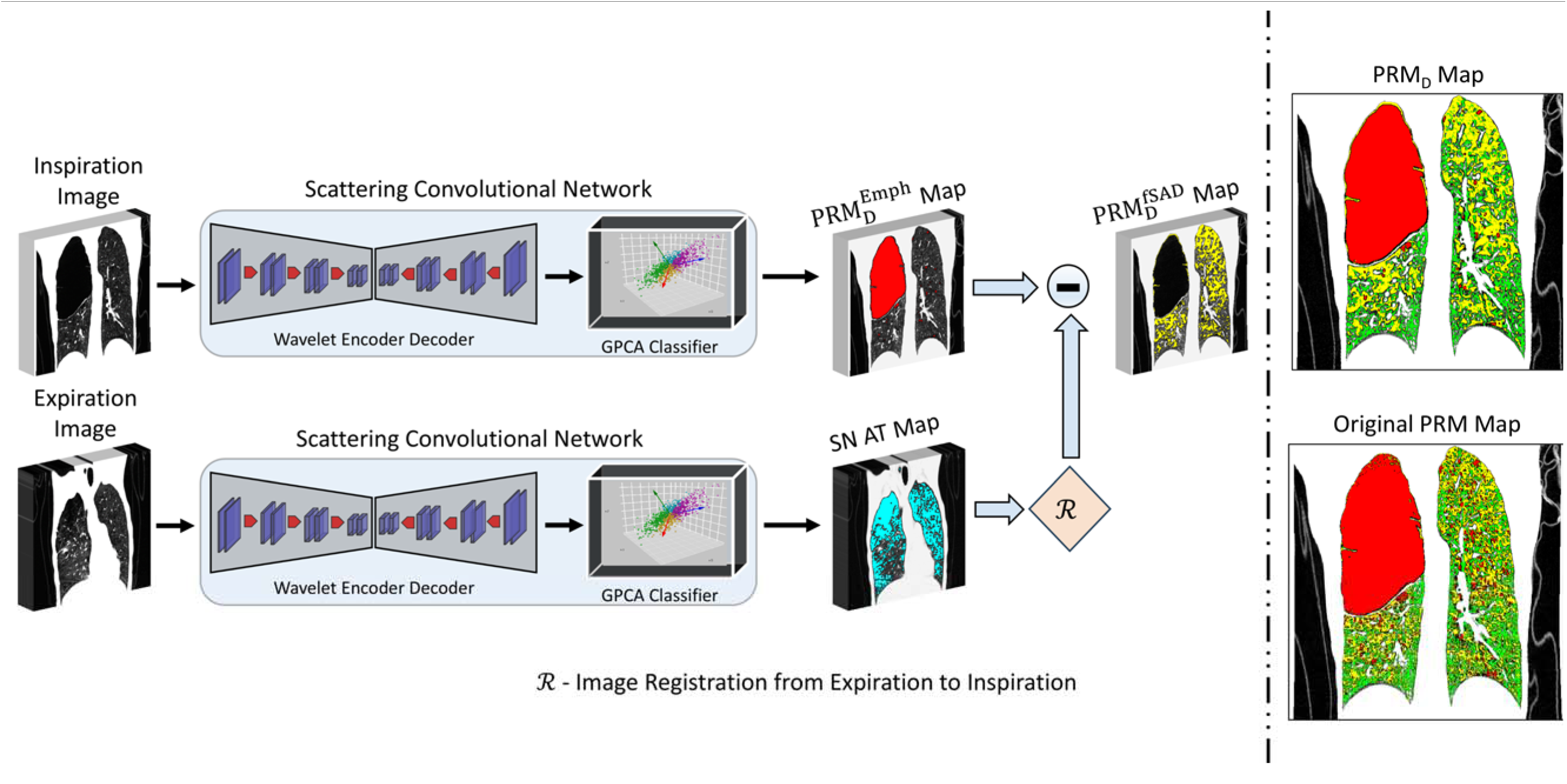
Workflow of feature-based Deep Parametric Response Mapping (PRM_D_) Paired inspiratory (I) and expiratory (E) CT volumes are processed by identical 3-D wavelet-scattering encoders that feed a generalized-PCA (GPCA) sub-space classifier. The inspiratory branch outputs an emphysema (Emph) map; the expiratory branch yields an air trapping (AT) map. After non-rigidly registering the AT map to the inspiratory space (:R.), AT voxels overlapping Emph are removed, producing the final PRM_D_-fSAD map, while PRM_D_-Emph is taken directly from the inspiratory classifier. The composite PRM_D_ (right, upper) is visually compared with the reference density-threshold PRM (right, lower). Red = emphysema, yellow = fSAD, green = normal lung.

### Model Training, Validation, and Testing

The PRM_D_ method was trained on a desktop workstation running a 64-bit Windows operating system (Windows 10) with an Intel Xeon W-2123 CPU at 3.6GHz with 128GB DDR6 RAM and an NVIDIA GeForce RTX 4080 graphic card with 2944 CUDA cores (NVIDIA driver 411.63) and 12GB GDDR RAM. Our proposed PRM_D_ was trained to learn the affine linear space that minimized the expected quadratic error between the scattering feature vector of the data and the expected feature vector over the signal class, for classification of different signal classes (i.e., emphysema and fSAD in our case) (18, 19). The x, y, and z-dimensions of the CT images in our dataset were x = 512, y = 512, z ∼ 600. We divided N = 8,972 subjects into 2,152 subjects (∼25%) training, 538 subjects (5%) validation, and 6,282 subjects (70%) testing sets to evaluate the performance of the PRM_D_ method. The training process involved optimizing the linear subspace dimension *d* and the invariance scale 2^J^. Emphysema and air trapping segmentation maps generated using standard density-based PRM were used for training the PRM_D_ method. We used a nested 3-fold cross-validation strategy for training the generalized PCA classifier in PRM_D_, where the outer loop was run ten times, and data were split into three equal pools internally. The generalized PCA classifier was trained on two pools and tested on the third. The scattering transform in the proposed PRM_D_ method was computed using the *Kymatio* package (20) implemented in PyTorch and run under the Python environment (version 3.10; Python software Foundation, Wilmington, Del; https://www.pyhon.org/). The generalized PCA classifier training and testing and the PRM_D_ method were developed in MATLAB version 9.13.0 (R2023a, Natick, Massachusetts: The MathWorks Inc.).

### Quantitative Assessment and Statistical Analysis

We evaluated PRM_D_ by comparing voxel-level classifications and emphysema and fSAD percentages to those from standard density-based PRM (reference standard). For each subject, we calculated the percentage of lung voxels classified as emphysema and fSAD by PRM_D_ and compared these to PRM values. Mean ± SD values were reported, and Pearson correlation assessed agreement between methods. Bland-Altman analysis (21) detected systematic bias by calculating mean differences and 95% limits of agreement. Voxel-level confusion matrices quantified agreement in labeling between PRM_D_ and PRM, from which Dice similarity coefficients (DSC) were derived for emphysema, fSAD, and normal classes. High DSC indicates similar lung partitioning. Performance was also analyzed in mild (GOLD 1–2) and severe (GOLD 3–4) COPD subgroups, with correlations and DSC recalculated and compared using two-sample tests.

To assess robustness, synthetic mixed Poisson-Gaussian noise (representing photon-counting and electronic noise (22)) was added at low, moderate, and high levels (with standard deviations σ ≈ 1X 10^−6^, 1X 10^−3^, 1X 10^−1^) to CT images in 2,200 test cases. PRM and PRM_D_ were run at each noise level, and changes in %emphysema and %fSAD relative to baseline were recorded. The average absolute error introduced by noise was calculated for each method, with paired t-tests evaluating whether PRM_D_ significantly reduced error. Continuous variables are reported as mean ± SD. Group differences were tested using Student’s t-tests or Mann-Whitney U tests. Statistical significance was defined as *p* < 0.05. All analyses were performed using MATLAB 9.13.0 (R2023a, The MathWorks Inc.).

## Results

### Deep PRM vs Standard PRM: Voxel Classification Agreement

The proposed PRM_D_ method achieved voxel classification and segmentations similar to standard PRM. Qualitatively, PRM_D_ maps of normal, fSAD, and emphysema regions were nearly indistinguishable from density-based PRM maps in most cases. Figure 2 shows representative GOLD 1 (mild) and GOLD 4 (severe) subjects in whom PRM_D_ and PRM outputs are overlaid; the spatial distribution of yellow (fSAD) and red (emphysema) regions matches closely between methods. Quantitatively, voxel-wise agreement was high (Table 2): overall, 95% of lung voxels across all test subjects received the same class label from PRM_D_ as from PRM. The class-specific Dice similarity coefficients were 0.96 for normal lung, 0.94 for fSAD, and 0.96 for emphysema, indicating excellent overlap.

**Figure 2.**
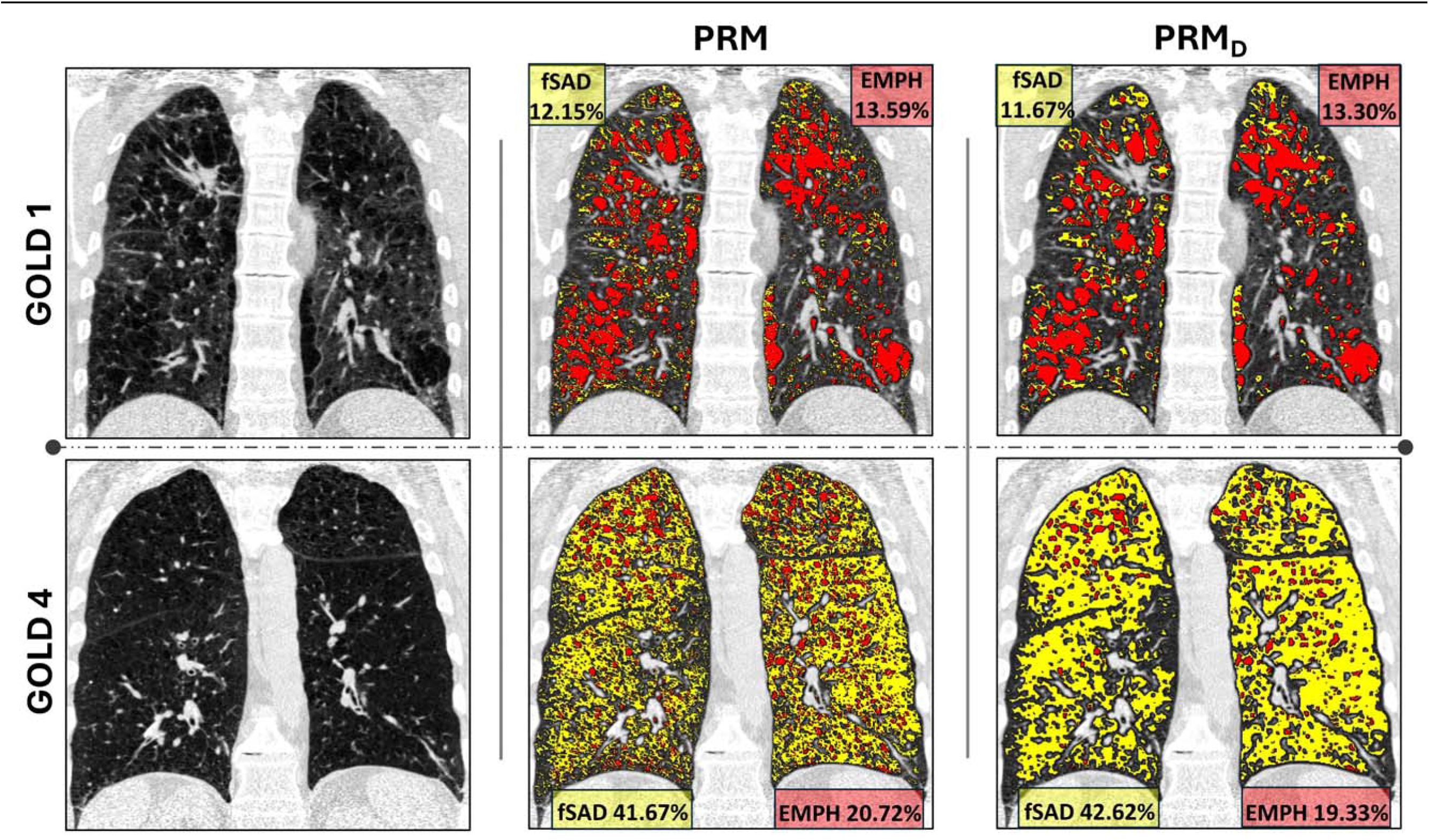
Visual comparison of PRM_D_ and standard PRM in mild and severe COPD. Coronal slices from an expiratory (EXP) CT scan (left column) with corresponding PRM (center) and PRM_D_ (right) overlays for (top row) GOLD 1 and (bottom row) GOLD 4 subjects. In each map, red denotes emphysema and yellow denotes fSAD; in-panel boxes give global percentages. Deep PRM reproduces both the spatial pattern and absolute burden of disease seen with the threshold-based method across severity extremes.

**Table 2.** Segmentation Performance Metrics Between PRM and PRM_D_ for the Test Data (N = 6282)

| METRIC | REGIONS/AREAS OF |  |  | OVERALL |
| --- | --- | --- | --- | --- |
|  | fSAD | Emphysema | NORMAL LUNG |  |
| **Dice Coefficient (%)** | 94.32 (1.94) | 96.15 (1.16) | 95.73 (0.87) | 95.36 (1.22) |
| **Jaccard Index** | 0.91 (0.12) | 0.93 (0.11) | 0.92 (0.07) | 0.92 (0.09) |
| **Precision** | 0.93 (0.13) | 0.92 (0.09) | 0.94 (0.11) | 0.93 (0.10) |
| **Recall** | 0.91 (0.18) | 0.94 (0.11) | 0.95 (0.09) | 0.94 (0.13) |
| **F<sub>1</sub>-Score** | 0.92 (0.14) | 0.93 (0.10) | 0.94 (0.11) | 0.93 (0.12) |
| **Rand Index** | 0.91 (0.15) | 0.96 (0.12) | 0.92 (0.12) | 0.93 (0.13) |

NOTE: All values expressed as mean (SD) except categorical variables expressed as N (%). SD = standard deviation.

For each subject, we compared the percentage of lung classified as emphysema by PRM_D_ vs PRM. The mean difference was near zero (PRM_D_ gave 0.1% higher emphysema on average, 95% CI of difference: –0.3% to +0.5%). The Pearson correlation between PRM_D_ and PRM emphysema was r = 0.991 (p<0.0001), and the Bland-Altman bias was –0.05%, with limits of agreement ±1.2%. In other words, across the entire range (0–60% emphysema), PRM_D_ results were essentially identical to standard PRM (within ∼1–2% lung volume). Similarly, for fSAD percentage, PRM_D_ vs PRM had r = 0.974 and negligible bias (mean difference +0.2%, limits ±1.8%). Figure 3 plots PRM_D_ vs PRM fSAD% for all subjects, demonstrating the tight agreement (points lying near the line of identity). Thus, on a per-subject basis, the PRM_D_ method replicated conventional PRM measurements with high fidelity.

**Figure 3.**
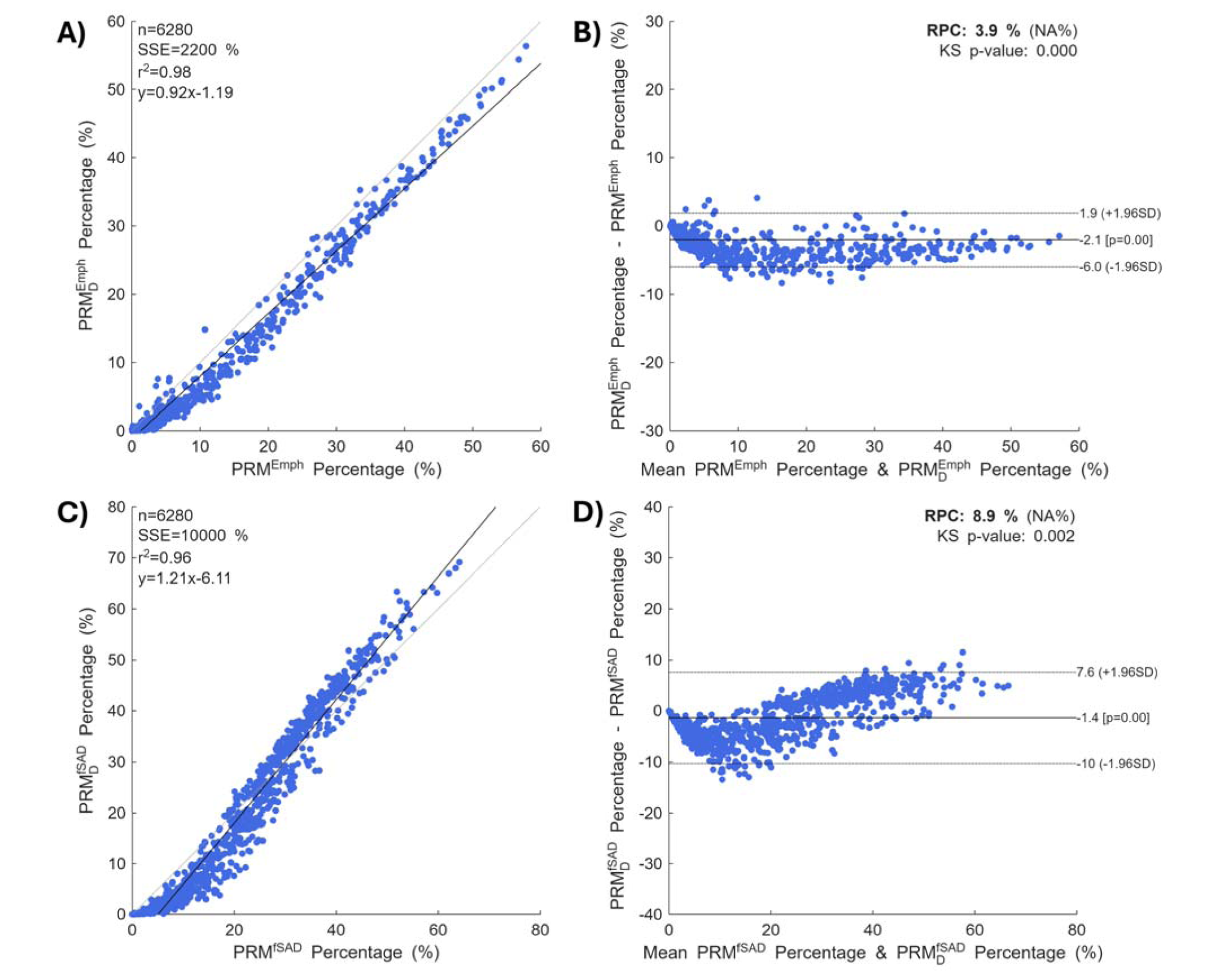
Agreement between PRM_D_ and standard PRM for whole-lung disease. A, B Emphysema: (A) correlation plot (identity line in grey) and (B) Bland-Altman analysis (mean bias –0.05%, limits ±1.2%). C, D fSAD: (C) correlation plot and (D) Bland-Altman plot (mean bias +0.2%, limits ±1.8%). SSE = sum-squared error; RPC = repeatability coefficient; KS = Kolmogorov–Smirnov test.

### Performance across GOLD stages

We examined whether agreement persists from normal spirometry to end-stage COPD. Over the full 6,282-subject test set, PRM_D_ versus density-based PRM yielded r² = 0.98 for emphysema (slope = 0.92; Figure 3A) and r² = 0.96 for fSAD (slope = 1.21; Figure 3C), corresponding to Pearson r ≈ 0.99 and 0.98. Bland–Altman plots (Figure 3B, D) showed negligible bias (–2.1% for emphysema, –1.4% for fSAD) with tight repeatability coefficients (RPC = 3.9% and 8.9%). Qualitative overlays in Figure 2 reinforce this finding: in both a mild (GOLD 1) and a severe (GOLD 4) exemplar, PRM_D_ reproduced the spatial distribution of fSAD (yellow) and emphysema (red) virtually identically to standard PRM. Stratifying by GOLD stage did not alter the quantitative relationship: bar charts in Figure 4 show that mean component volumes derived from PRM_D_ (panel A) overlap those from standard PRM (panel B) at every stage, with error bars fully super-imposed. Even in GOLD 4, where emphysema can exceed 50% of lung volume, PRM_D_ differed from the reference by no more than 2–3 absolute percentage points. Together, qualitative (Figure 2) and quantitative (Figures 3–4) evidence confirm that PRM_D_ mirrors conventional PRM with high fidelity throughout the full spectrum of COPD severity.

**Figure 4.**
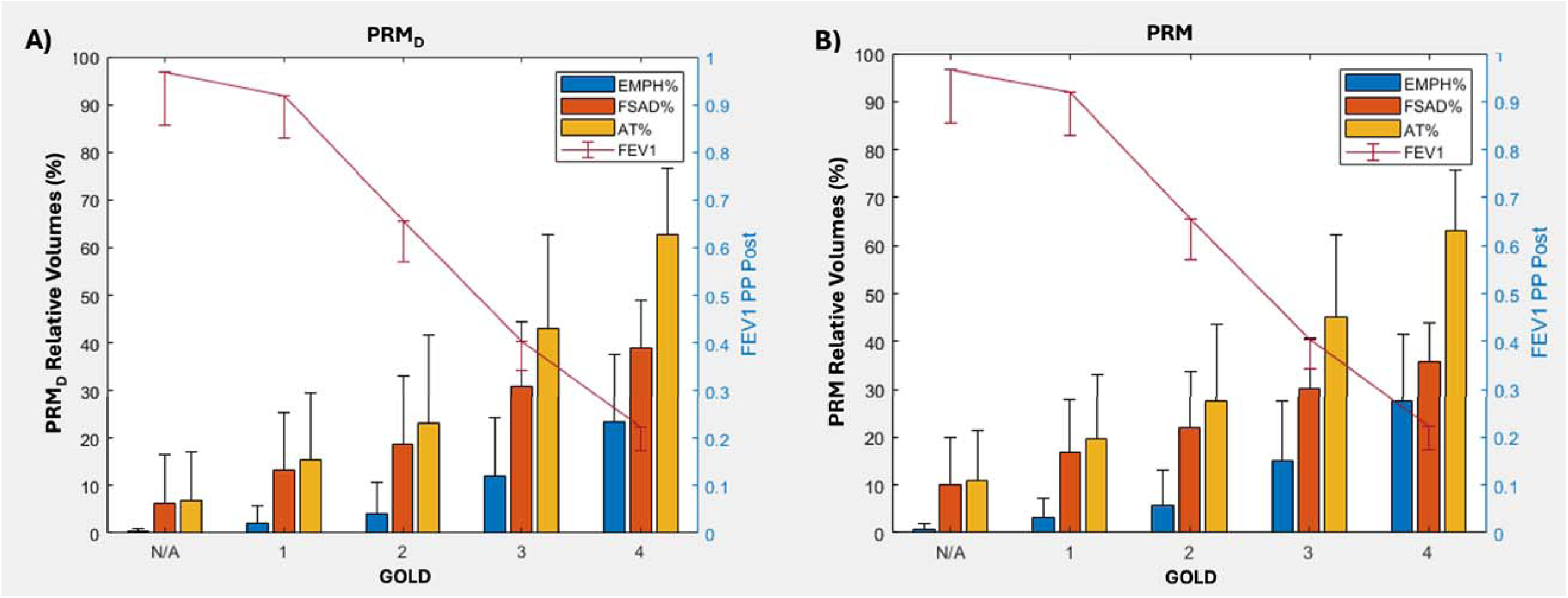
Relative volumes of emphysema, fSAD and total air trapping across GOLD stages. Bar graphs show mean ± SD percentages derived from (A) PRM_D_ and (B) density-based PRM. Superimposed red line depicts mean post-bronchodilator FEV_1_% predicted (right y-axis). PRM_D_ mirrors the trajectory of disease components and lung function decline observed with standard PRM.

### Correlation with Pulmonary Function

Both standard density-based PRM and the proposed PRM_D_ method showed statistically significant correlations with clinical, physiologic, and functional measures of COPD severity (Table 3). PRM_D_ demonstrated consistently stronger correlations across all parameters. For emphysema, PRM_D_ showed greater inverse associations with FEV (r = –0.54 vs –0.42; *P* < .0001) and FEV /FVC (r = –0.61 vs –0.54; *P* < .0001) compared to standard PRM. Similarly, PRM_D_-based fSAD measurements correlated more strongly with FEV (r = –0.51 vs –0.42; *P* < .0001) and FEV /FVC (r = –0.52 vs –0.45; *P* < .0001). PRM_D_ also improved associations with markers of hyperinflation (TLC: r = 0.65–0.70 vs 0.61–0.64) and physical function (6-minute walk distance: r = –0.55 to –0.62 vs –0.47 to –0.57). Quality-of-life and disease burden metrics, including the St. George’s Respiratory Questionnaire (r = 0.63–0.73) and BODE index (r = 0.60–0.62), similarly showed stronger correlations with PRM_D_ than standard PRM.

**Table 3.** Correlation of PRM and PRM_D_ with Clinical and Patient Assessment Parameters (N = 6282)

| CLINICAL PARAMETER | $PRM^{fSAD}$ | $PRM_D^{fSAD}$ | $PRM^{Emph}$ | $PRM_D^{Emph}$ |
| --- | --- | --- | --- | --- |
| **FEV<sub>1</sub>** | -0.42 (<0.0001) | -0.51 (<0.0001) | -0.42 (<0.0001) | -0.54 (<0.0001) |
| **FEV<sub>1</sub>/FVC** | -0.45 (<0.0001) | -0.52 (<0.0001) | -0.54 (<0.0001) | -0.61 (<0.0001) |
| **TLC** | 0.61 (0.03) | 0.65 (0.001) | 0.64 (<0.0001) | 0.70 (<0.0001) |
| **6-min Walk Distance (m)** | -0.47 (0.05) | -0.55 (0.001) | -0.57 (<0.0001) | -0.62 (<0.0001) |
| **SGRQ Score** | 0.62 (<0.0001) | 0.63 (<0.0001) | 0.71 (<0.0001) | 0.73 (<0.0001) |
| **BODE Score** | 0.53 (<0.0001) | 0.60 (<0.0001) | 0.56 (<0.0001) | 0.62 (<0.0001) |

NOTE: Presented are the $r$ ( $p$ ) values generated from Pearson correlation for each of the prognostic indicators when assessing the contribution of fSAD and emphysema defined by both PRM and PRM<sub>D</sub>. FEV<sub>1</sub> = forced expiratory volume in the first second, FVC = forced vital capacity, TLC = total lung capacity, SGRQ = St. George's Respiratory Questionnaire, BODE = body mass index, degree of airflow obstruction, dyspnea, and exercise capacity (as measure in a 6-minute walk test).

### Robustness to Noise and Image Misregistration

Gaussian noise experiments (σ = 10□□→1; Figure 5) showed that PRM_D_ sustains markedly higher voxel-level accuracy than density threshold-based standard PRM as image noise increases. For fSAD, Dice overlap with PRM_D_ stayed above 90% until σ ≈ 10□^6^ and remained about 60% at σ = 1, whereas for standard PRM it dropped from roughly 82% to 35%. Emphysema followed the same trend (PRM_D_ 92%→68% vs standard PRM 80%→50%). Slice overlays in Figure 5C highlight the practical impact: when low noise was added (σ = 10□□) conventional PRM inflated fSAD from 3.3% to 7.2% and emphysema from 2.8% to 8.0%, while PRM_D_ changed each metric by less than 2%. At σ = 10□³ the discrepancy widened—standard PRM nearly tripled both components (∼20%), whereas PRM_D_ limited the increase to about 7%.

**Figure 5.**
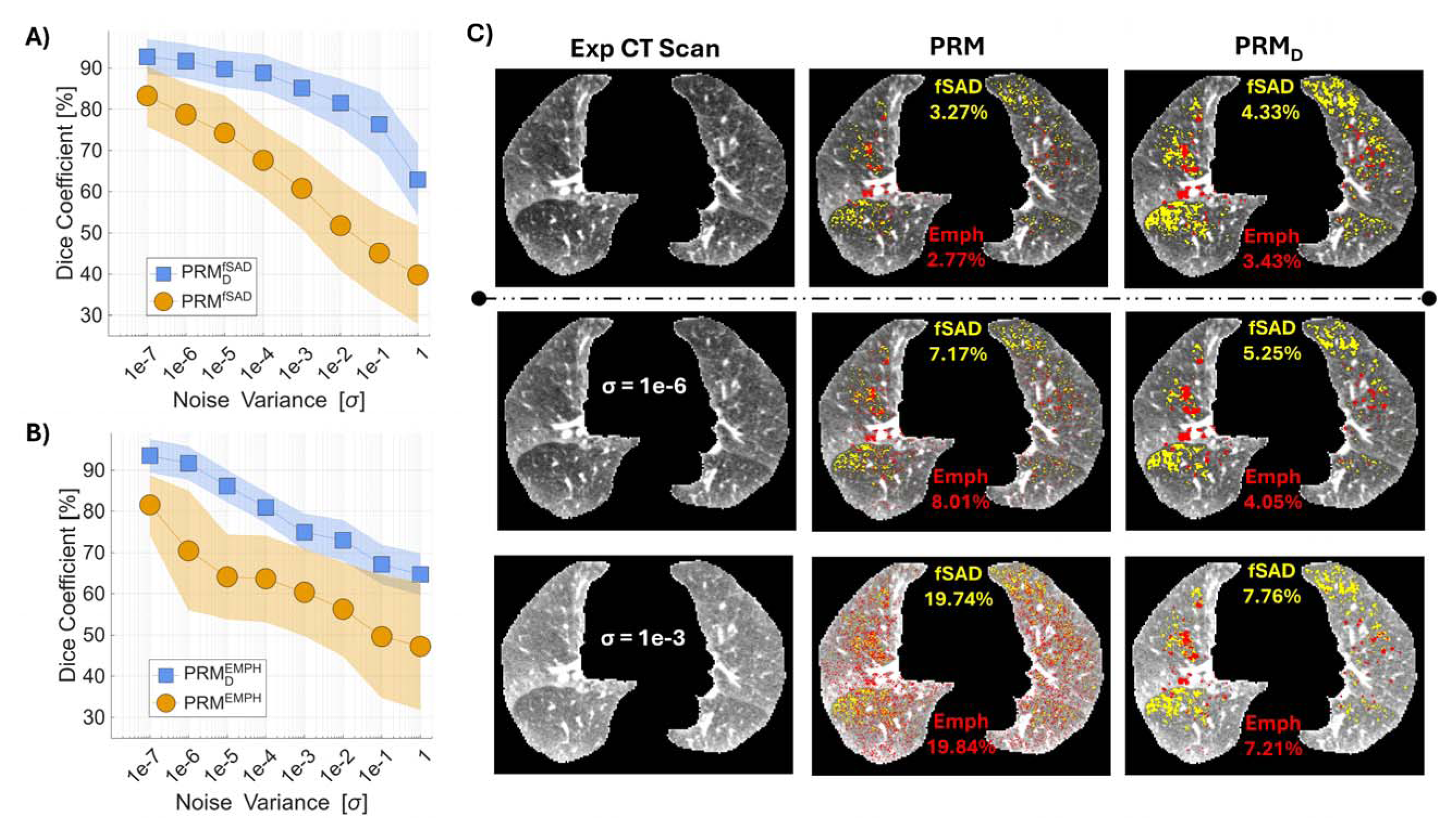
Noise-robustness analysis. (A) Dice coefficient for fSAD maps and (B) Dice for emphysema maps as a function of added mixed Poisson–Gaussian noise variance (σ). Shaded bands = ±SD. (C) Representative axial expiratory CT slices (left) and corresponding PRM vs PRM_D_ overlays at baseline, σ = 1 × 10^−6^ and σ = 1 × 10^−3^. PRM_D_ retains higher overlap with the noise-free reference, limiting over-segmentation (yellow/red) relative to standard PRM.

We also analyzed sensitivity to residual inspiration–expiration mis-registration. Recomputing PRM_D_ emphysema after an explicit non-rigid registration step produced r² = 1.00, bias = 0.01% and limits of agreement ± 0.55% (Figure 6A, B); visual comparison (Figure 6C) showed negligible differences.

**Figure 6.**
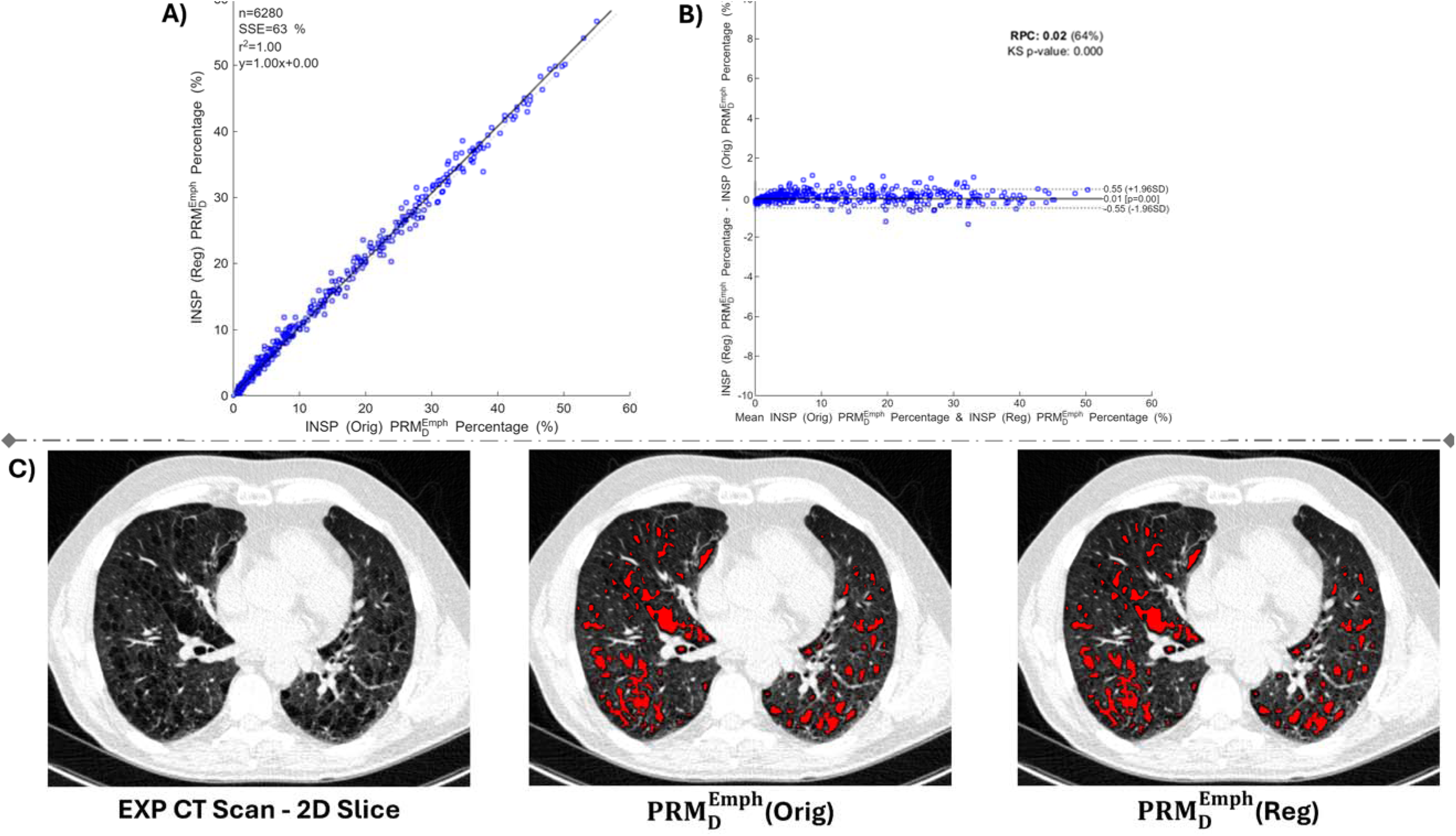
Influence of residual inspiratory–expiratory mis-registration on PRM_D_ emphysema quantification. (A) Correlation and (B) Bland-Altman plots comparing original PRM_D_ Emph% to values after explicit non-rigid registration (Reg). Agreement is essentially perfect (r² = 1.00; bias 0.01%, limits ±0.55%). (C) Axial expiratory CT slice with emphysema masks from original and registered PRM_D_ demonstrating negligible visual differences.

## Discussion

Deep Parametric Response Mapping (PRM_D_) is an approach to COPD phenotyping that combines wavelet scattering transform features with subspace learning to classify lung voxels on paired inspiratory–expiratory CT scans. PRM_D_ closely replicates conventional density-based PRM classifications for emphysema and functional small airways disease (fSAD), achieving 95% voxel-wise agreement and r = 0.98 correlation with standard PRM metrics. By replacing thresholding with feature-based classification, PRM_D_ offers improved resilience to image noise and better captures complex tissue characteristics beyond intensity cutoffs.

Standard PRM has been widely used to differentiate emphysema from air trapping in COPD and serves as a foundation for evaluating longitudinal progression and treatment effects (2, 22–25). In our implementation, PRM_D_ learned to replicate standard PRM using fixed threshold labels (–950 HU for emphysema, –856 HU for air trapping), but relied instead on mathematically defined scattering filters that extract translation-invariant features. This approach captures local textural and spatial patterns relevant to disease, reducing dependence on scanner-and dose-specific intensity variations.

A key strength of PRM_D_ lies in its robustness to imaging noise and variability. Under simulated noise conditions, standard PRM overestimated emphysema and fSAD by approximately 15%, while PRM_D_ limited these errors to under 5% (*P* < .001). These improvements are clinically meaningful, especially in longitudinal studies or multi-center trials where repeated scans may vary in quality due to inspiratory effort, scanner differences, or protocol inconsistencies. PRM_D_’s stability reduces the likelihood of false changes in imaging biomarkers and may improve sensitivity to true biological progression. Importantly, PRM_D_ preserved—and in many cases strengthened—the correlations between imaging phenotypes and clinical metrics. PRM_D_-derived emphysema and fSAD showed stronger associations with FEV , FEV /FVC, six-minute walk distance, St. George’s Respiratory Questionnaire (SGRQ) score, and BODE index compared to standard PRM. These results suggest that PRM_D_ provides a more physiologically relevant measure of disease burden and may be better suited for precision imaging applications in COPD.

Our work aligns with and extends prior studies applying deep learning to improve quantification of air trapping and emphysema (26, 27). Unlike approaches using personalized thresholding or synthetic expiratory image generation via GANs (28, 29), PRM_D_ directly uses real paired inspiratory and expiratory CT scans, improving interpretability and avoiding potential biases introduced by synthetic data. Furthermore, the wavelet scattering transform offers theoretical stability guarantees and requires less training data than typical convolutional networks, making it practical for clinical datasets. The success of this architecture in replicating PRM underscores the value of handcrafted, fixed filters for radiologic tasks.

This study has several limitations. First, PRM_D_ was validated using standard PRM as ground truth, since voxel-level histopathologic correlation is not widely available. While practical, this approach assumes PRM is accurate and may overlook its own thresholding limitations. Second, the model used three spatial scales, 12 orientations, and second-order scattering coefficients; incorporating higher-order features or finer angular resolution may enhance classification. Lastly, we tested a single Poisson-Gaussian noise model. Real-world artifacts such as motion blur or beam hardening were not simulated, and future work should assess PRM_D_ performance under broader acquisition variability.

## Conclusion

This study demonstrates that PRM_D_—a combination of wavelet scattering networks and subspace learning—accurately replicates conventional PRM assessments while enhancing robustness to noise. By eliminating manual thresholding and using interpretable, fixed filters, PRM_D_ maintains strong correlations with lung function and symptom burden, supporting its clinical relevance. This feature-driven method reduces variability in quantitative CT analysis and improves sensitivity to true disease changes. Future research should focus on validating PRM_D_’s longitudinal performance and applying it in clinical trials where reliable quantification of emphysema and fSAD is essential for assessing treatment efficacy.

## Data Availability

The datasets described in this study are not readily available because they are part of an NIH sponsored clinical trial and require a data use agreement to be signed. For access to COPDGene data visit https://www.copdgene.org/phase-1-study-documents.htm for instructions.

## Abbreviations

PRM_D_: Deep Parametric Response Mapping
fSAD: Functional Small Airways Disease
CNN: Convolutional Neural Network
PRISm: Preserved Ratio Impaired Spirometry
FEV_1_: forced expiratory volume in the first second
FVC: forced vital capacity
TLC: total lung capacity measured on volumetric computed tomography of the chest at maximal inhalation
FRC: functional residual capacity measured on volumetric computed tomography of the chest at the end of tidal exhalation
LAA: low attenuation areas,
Pi10: square root of the wall area of a theoretical airway of 10 mm luminal perimeter

## Acknowledgments

This work was supported by Department of Defense grant HT94252310698 and National Heart, Blood, and Lung Institute grants R01HL162661, U01 HL089897 and U01 HL089856 and by National Institutes of Health contract 75N92023D00011. The COPDGene study (NCT00608764) has also been supported by the COPD Foundation through contributions made to an Industry Advisory Committee that has included AstraZeneca, Bayer Pharmaceuticals, Boehringer-Ingelheim, Genentech, GlaxoSmithKline, Novartis, Pfizer, and Sunovion.

## References

1. Han MK, Agusti A, Calverley PM, Celli BR, Criner G, Curtis JL, et al. Chronic obstructive pulmonary disease phenotypes: the future of COPD. Am J Respir Crit Care Med. 2010;182(5):598–604.

2. Galbán CJ, Han MK, Boes JL, Chughtai KA, Meyer CR, Johnson TD, et al. Computed tomography-based biomarker provides unique signature for diagnosis of COPD phenotypes and disease progression. Nat Med. 2012;18(11):1711–5.

3. Labaki WW, Gu T, Murray S, Hatt CR, Galban CJ, Ross BD, et al. Reprint of: Voxel-Wise Longitudinal Parametric Response Mapping Analysis of Chest Computed Tomography in Smokers. Acad Radiol. 2019;26(3):306–12.

4. Boes JL, Bule M, Hoff BA, Chamberlain R, Lynch DA, Stojanovska J, et al. The Impact of Sources of Variability on Parametric Response Mapping of Lung CT Scans. Tomography. 2015;1(1):69–77.

5. Goris ML, Zhu HJ, Blankenberg F, Chan F, Robinson TE. An automated approach to quantitative air trapping measurements in mild cystic fibrosis. Chest. 2003;123(5):1655–63.

6. Ram S, Hoff BA, Bell AJ, Galban S, Fortuna AB, Weinheimer O, et al. Improved detection of air trapping on expiratory computed tomography using deep learning. PLoS One. 2021;16(3):e0248902.

7. Shi J, Zhao Y, Xiang W, Monga V, Liu X, Tao R. Deep scattering network with fractional wavelet transform. IEEE Transactions on Signal Processing. 2021;69:4740–57.

8. Liu L, Wu J, Li D, Senhadji L, Shu H. Fractional wavelet scattering network and applications. IEEE Transactions on Biomedical Engineering. 2018;66(2):553–63.

9. Regan EA, Hokanson JE, Murphy JR, Make B, Lynch DA, Beaty TH, et al. Genetic epidemiology of COPD (COPDGene) study design. Copd. 2010;7(1):32–43.

10. Agusti A, Celli BR, Criner GJ, Halpin D, Anzueto A, Barnes P, et al. Global Initiative for Chronic Obstructive Lung Disease 2023 Report: GOLD Executive Summary. Arch Bronconeumol. 2023;59(4):232–48.

11. Belloli EA, Degtiar I, Wang X, Yanik GA, Stuckey LJ, Verleden SE, et al. Parametric Response Mapping as an Imaging Biomarker in Lung Transplant Recipients. Am J Respir Crit Care Med. 2017;195(7):942–52.

12. Bhatt SP. Imaging Small Airway Disease: Probabilities and Possibilities. Ann Am Thorac Soc. 2019;16(8):975–7.

13. Young RK. Wavelet theory and its applications: Springer Science & Business Media; 2012.

14. Razali NF, Isa IS, Sulaiman SN, Karim NKA, Osman MK. CNN-Wavelet scattering textural feature fusion for classifying breast tissue in mammograms. Biomedical Signal Processing and Control. 2023;83:104683.

15. Jin Y, Duan Y. Wavelet scattering network-based machine learning for ground penetrating radar imaging: Application in pipeline identification. Remote Sensing. 2020;12(21):3655.

16. Ram S, Tang W, Bell AJ, Pal R, Spencer C, Buschhaus A, et al. Lung cancer lesion detection in histopathology images using graph-based sparse PCA network. Neoplasia. 2023;42:100911.

17. Ram S. Sparse representations and nonlinear image processing for inverse imaging solutions: The University of Arizona; 2017.

18. Bruna J, Mallat S. Invariant scattering convolution networks. IEEE Trans Pattern Anal Mach Intell. 2013;35(8):1872–86.

19. Ma Y, Derksen H, Hong W. Segmentation of multivariate mixed data via Lossy data coding and compression. IEEE Trans Pattern Anal Mach Intell. 2007;29(9):1546–62.

20. Mathieu Andreux TA, Georgios Exarchakis, Roberto Leonarduzzi, Gaspar Rochette, Louis Thiry, John Zarka, Stéphane Mallat, Joakim Andén, Eugene Belilovsky, Joan Bruna, Vincent Lostanlen, Muawiz Chaudhary, Matthew J. Hirn, Edouard Oyallon, Sixin Zhang, Carmine Cella, Michael Eickenberg. Kymatio: Scattering Transforms in Python. Journal of Machine Learning Research. 2020;21(60):1–6.

21. Giavarina D. Understanding bland altman analysis. Biochemia medica. 2015;25(2):141–51.

22. Foi A, Trimeche M, Katkovnik V, Egiazarian K. Practical Poissonian-Gaussian noise modeling and fitting for single-image raw-data. IEEE Trans Image Process. 2008;17(10):1737–54.

23. Pompe E, Galbán CJ, Ross BD, Koenderman L, Ten Hacken NH, Postma DS, et al. Parametric response mapping on chest computed tomography associates with clinical and functional parameters in chronic obstructive pulmonary disease. Respir Med. 2017;123:48–55.

24. Boes JL, Hoff BA, Bule M, Johnson TD, Rehemtulla A, Chamberlain R, et al. Parametric Response Mapping Monitors Temporal Changes on Lung CT Scans in the Subpopulations and Intermediate Outcome Measures in COPD Study (SPIROMICS). Academic Radiology. 2015;22(2):186–94.

25. Capaldi DP, Zha N, Guo F, Pike D, McCormack DG, Kirby M, et al. Pulmonary Imaging Biomarkers of Gas Trapping and Emphysema in COPD: (3)He MR Imaging and CT Parametric Response Maps. Radiology. 2016;279(2):597–608.

26. Hasenstab KA, Tabalon J, Yuan N, Retson T, Hsiao A. CNN-based Deformable Registration Facilitates Fast and Accurate Air Trapping Measurements at Inspiratory and Expiratory CT. Radiology: Artificial Intelligence. 2022;4(1):e210211.

27. Kim JH, Yoon HJ, Lee E, Kim I, Cha YK, Bak SH. Validation of Deep-Learning Image Reconstruction for Low-Dose Chest Computed Tomography Scan: Emphasis on Image Quality and Noise. Korean J Radiol. 2021;22(1):131–8.

28. Chen B, Liu Z, Lu J, Li Z, Kuang K, Yang J, et al. Deep learning parametric response mapping from inspiratory chest CT scans: a new approach for small airway disease screening. Respir Res. 2023;24(1):299.

29. Chaudhary MFA, Awan HA, Gerard SE, Bodduluri S, Comellas AP, Barjaktarevic I, et al. Deep Learning Estimation of Small Airways Disease from Inspiratory Chest CT: Clinical Validation, Repeatability, and Associations with Adverse Clinical Outcomes in COPD. Am J Respir Crit Care Med. 2025.

